# Early estimation of the risk factors for hospitalisation and mortality by COVID-19 in México

**DOI:** 10.1101/2020.05.11.20098145

**Authors:** María Fernanda Carrillo-Vega, Guillermo Salinas-Escudero, Carmen García-Peña, Luis Miguel Gutiérrez-Robledo, Lorena Parra-Rodríguez

## Abstract

**Background:** With its high prevalence of chronic non-degenerative diseases, it is suspected that in Mexico there is a high risk of fatal complications from COVID-19. The present study aims to estimate the risk factors for hospitalisation and death in the Mexican population infected by SARS-CoV-2.

**Methods and Findings:** We used the publicly available data released by the Epidemiological Surveillance System for Viral Respiratory Diseases of the Mexican Ministry of Health (Secretaría de Salud, SS). All records of positive SARS-CoV-2 cases were included. Two multiple logistic regression models were fitted to estimate the association between the hospitalisation and mortality, with other covariables. Data on 10,544 individuals (57.68% men), with mean age 46.47±15.62, were analysed. Men were about 1.54 times as likely to be hospitalized than women (p<0.001, 95% C.I. 1.37-1.74); individuals aged 50-74 and ≥74 years were more likely to be hospitalized than people from 25-49 years (OR 2.05, p<0.001, 95% C.I. 1.81-2.32, and OR 23.84, p<0.001, 95% C.I. 2.90-5.15, respectively). People with hypertension, obesity, and diabetes were more likely to be hospitalised than people without these morbidities (p<0.01). Men had more risk of death in comparison to women (OR=1.53, p<0.001, 95% C.I. 1.30-1.81) and individuals aged 50-74 and ≥75 years were more likely to die than people from 25-49 years (OR 1.96, p<0.001, 95% C.I. 1.63-2.34, and OR 3.74, p<0.001, 95% C.I. 2.80-4.98, respectively). Hypertension, obesity, and diabetes presented in combination, provided a higher risk of dying in comparison to not having these diseases (OR=2.10; p<0.001, 95% C.I. 1.50-2.93). Hospitalisation, intubation and pneumonia conferred a higher risk of dying (OR 5.02, p<0.001, 95% C.I. 3.88-6.50; OR 4.27, p<0.001, 95% C.I. 3.26-5.59, and OR=2.57; p<0.001, 95% C.I. 2.11-3.13, respectively). The main limitation of our study is the lack of information on mild (asymptomatic) or moderate cases of COVID-19.

**Conclusions:** The present study points out that in Mexico, where an important proportion of the population develops two or more chronic conditions simultaneously, high mortality is a sever outcome for those infected by SARS-CoV-2.

## INTRODUCTION

Since the start of the epidemic of the COVID-19 disease in Wuhan, China in December 2019, the SARS-CoV-2 has continued to spread globally, resulting in more than 3 million confirmed cases and about 230 thousand deaths around the globe by the first week of May 2020 (1). The up-to-date data evidence that the growth of the pandemic had slowed in Asia, while Europe and America contribute to the highest number of cases worldwide. The last is the region with the highest incidence, contributing 49% of cases across the World Health Organization (WHO) regions in the last 14 days of April. Since the first case reported in America by The United States on January 31^th^, the SARS-CoV-2 disease has spread to 50 countries and territories in the region, including Mexico (2). This country confirmed the first case of COVID-19 on February 27^th^ and the first death on March 19^th^. On March 23^th^ the health authorities declared the COVID-19 as a health emergency, starting the social isolation as the main action to contain the epidemic (3). The acceleration phase in Mexico was declared almost a month later, on April 21^st^, when the number of confirmed cases was 9,501 and 857 total deaths had been registered.

The first information on the COVID-19 was generated in China, where the disease transmission mechanisms, the incubation period and the clinical manifestations had been described. (4-7). It has been stated that most of the infected individuals recover spontaneously before approximately 7 to 10 days, while the rest, on the other hand, develop fatal complications including organ failure, septic shock, pulmonary edema, severe pneumonia, and Acute Respiratory Distress Syndrome (ARDS) (5). Older persons and those with underlying health conditions are at higher risk of severe disease and death (8).

Although the aging rate is not as high as in other countries, Mexico has one of the highest prevalence of chronic non-degenerative diseases, especially obesity, diabetes, and hypertension, in both young and older adults (9). In this respect, it is suspected that the risk of fatal complications is higher than in other countries. Amidst the global health emergency and considering that the knowledge of such information is crucial for the government and the health authorities in the decision-making process during the epidemic, this work aims to estimate the risk factors for hospitalisation and death in the Mexican population infected by SARS-CoV-2.

## METHODS

The present analysis was based on the publicly available data of the suspected cases of viral respiratory disease released by the Epidemiological Surveillance System for Viral Respiratory Diseases of the Mexican Ministry of Health (Secretaría de Salud, SS) on April 23rd, 2020 (10). This system contains data collected by 475 viral respiratory disease monitoring units (USMER) located in the different health services throughout the country, and directly from the medical units that attended the cases.

The dataset includes positive, negative, and suspected cases to COVID, with or without pneumonia, both in ambulatory and hospital management. Information on sex, age, nationality, residence place, and migratory status had been registered. Information on the type of medical unit of first contact (USMER or not USMER) and if the individual was hospitalized or stayed in outpatient management, in public or private services was included. The dates when the individual developed the COVID-19 symptoms, were admitted to hospitalisation and death were also recorded. Comorbidities encompassed hypertension, diabetes, obesity, cardiovascular disease, chronic obstructive pulmonary disease (COPD), asthma, chronic kidney disease (CKD), immunosuppression, and other diseases reported by the individual. Other risk factors for severe COVID-19 included smoking and pregnancy. The database does not contain evolution during the stay in the medical units.

For the present analysis, all records of positive SARS-CoV-2 cases were included. No record was deleted due to the presence of missing data since the number of missing values was <2% of the total registers. The data cutoff for the study was April 23rd, 2020.

### Variables

Two primary endpoints were defined: hospitalisation and death. The former defined as individuals who tested positive for COVID-19 and required hospitalisation. The last was defined as positive cases for COVID-19 who died, regardless of being hospitalized or remained in outpatient management.

Sex, morbidity, pregnancy, immune-suppression condition, smoking, pneumonia, intubation, admission to ICU, days from the presentation of symptoms to the hospitalization and to death, and from admission to health care unit to death, as well as the health service that provided the treatment, are reported in the present work.

Because of the high prevalence of chronic diseases in Mexico, especially hypertension, diabetes, and obesity, in addition to reporting its prevalence alone, these morbidities were grouped to determine whether together impose a different risk.

### Statistical analysis

Descriptive analysis was performed. Continuous variables are presented as means and standard deviations, and categorical variables are expressed as number and percentage. Comparisons of hospitalized vs. no hospitalized individuals and survivors vs. no survivors were estimated through the Mann-Whitney test for continuous variables and χ^2^ for categorical variables. From this point, only individuals over 25 years were included in the analysis, as in the above group only 5 deaths had been registered.

Two multiple logistic regression models were fitted to estimate the association between the hospitalisation and mortality, with the rest of the covariables. Those variables that resulted not significant were excluded from the final model. All analyses were performed with the statistical package software STATA 14.

## RESULTS

Data on 10,544 individuals with mean age 46.47±15.62 were analysed. From this total, 57.68% (n=6,082) were men (Table 1). Hypertension was the most prevalent morbidity (21.74%), followed by obesity (20.05%) and diabetes (17.65%). It can be seen that the prevalence and number of morbidities increased as age did. From the general population, 55.61% (n=5797) had no comorbidities. The most frequent combination of chronic diseases was diabetes and hypertension (6.05%). This combination is followed by hypertension and obesity (3.75%), Diabetes and Hypertension and Obesity (3.06%), and diabetes and obesity (2.2%). Obesity in combination with hypertension (5.41%) and diabetes (3.01%) was more frequent in individuals from 50-74 years. Obesity alone was more frequent (13.4%) in the 25-49 age group. From the total number of COVID-19 cases, 28.68% developed pneumonia, that was more frequent in individuals over 50 years (61.14% of the total cases), and 37.91 % were hospitalised, mainly individuals over 50 years (59.79%). Hospitalisation, intubation, and intensive care unit admission were more frequent in the older age groups. From the total individuals 968 (9.18%) died. It can be seen that deaths increased as age did, with the higher percentage in the older age group. Death was presented in 9.18% of the total number of cases, 0.84% in individuals under 25 years, 4.52% in the 25-49 years group, 14.99% in people between 50 and 74, and 27.95% for the older age group. The mean of days from the symptomatology presentation to the admission was 4.32±3.39 days, from the admission to the dead 5.88±4.95 days, and from the symptomatology presentation to the dead 10.08±5.47 days. The health ministry services (SSA, 48.44%) and the Mexican Institute of Social Services (IMSS, 34.47%) were the institutions that attended the largest number of cases (Table 1).

**Table 1.** General characteristics by age group

|  | **Total**<br>**n = 10544** | **<25 y**<br>**n= 598** | **25-49 y**<br>**n= 5640** | **50-74 y**<br>**n= 3823** | **≥75 y**<br>**n= 483** |
| --- | --- | --- | --- | --- | --- |
| Sex (men) | 6082 (57.68) | 329 (55.02) | 3203 (56.79) | 2286 (59.8) | 264 (54.66) |
| Hypertension | 2272 (21.74) | 7 (1.17) | 622 (11.11) | 1372 (36.31) | 271 (56.58) |
| Obesity | 2097 (20.05) | 44 (7.37) | 1115 (19.91) | 864 (22.84) | 74 (15.48) |
| Diabetes | 1845 (17.65) | 11 (1.84) | 525 (9.38) | 1148 (30.38) | 161 (33.68) |
| Cardiovascular Disease | 312 (2.99) | 2 (0.34) | 57 (1.02) | 197 (5.22) | 56 (11.69) |
| Chronic kidney disease | 222 (2.13) | 3 (0.5) | 61 (1.09) | 135 (3.58) | 23 (4.81) |
| COPD | 271 (2.59) | 1 (0.17) | 39 (0.7) | 156 (4.13) | 75 (15.66) |
| Asthma | 374 (3.58) | 18 (3.02) | 243 (4.34) | 101 (2.68) | 12 (2.51) |
| Number of Comorbidities | 0.7 ± 0.96 | 0.14 ± 0.42 | 0.47 ± 0.77 | 1.04 ± 1.08 | 1.4 ± 1.15 |
| No Comorbidities | 5797 (55.61) | 524 (88.07) | 3692 (66.05) | 1465 (38.91) | 116 (24.42) |
| Combinated Chronic Diseases |  |  |  |  |  |
| Diabetes & Hypertension & Obesity | 323 (3.06) | 1 (0.17) | 86 (1.52) | 207 (5.41) | 29 (6.0) |
| Diabetes & Hypertension | 638 (6.05) | 0 | 115 (2.04) | 440 (11.51) | 83 (17.18) |
| Diabetes & Obesity | 232 (2.2) | 4 (0.67) | 109 (1.93) | 115 (3.01) | 4 (0.83) |
| Hypertension & Obesity | 395 (3.75) | 0 | 163 (2.89) | 207 (5.41) | 25 (5.18) |
| Only Hypertension | 914 (8.67) | 6 (1.0) | 258 (4.57) | 517 (13.52) | 133 (27.54) |
| Only Obesity | 1142 (10.83) | 39 (6.52) | 756 (13.4) | 331 (8.66) | 16 (3.31) |
| Only Diabetes | 649 (6.16) | 6 (1.0) | 215 (3.81) | 384 (10.04) | 44 (9.11) |
| Pregnancy | 54 (0.51) | 6 (1.01) | 48 (0.85) | -- | -- |
| Immuno-suppressed | 207 (1.98) | 12 (2.01) | 76 (1.36) | 99 (2.62) | 20 (4.18) |
| Smoking | 936 (8.96) | 42 (7.04) | 517 (9.24) | 322 (8.53) | 55 (11.48) |
| Pneumonia | 3024 (28.68) | 53 (8.86) | 1122 (19.89) | 1581 (41.35) | 268 (55.49) |
| Hospitalized | 3997 (37.91) | 75 (12.54) | 1532 (27.16) | 2040 (53.36) | 350 (72.46) |
| Intubated | 466 (4.42) | 6 (1.01) | 123 (2.18) | 289 (7.56) | 48 (9.94) |
| ICU | 492 (4.67) | 6 (1.01) | 153 (2.71) | 296 (7.74) | 37 (7.66) |
| Dead | 968 (9.18) | 5 (0.84) | 255 (4.52) | 573 (14.99) | 135 (27.95) |
| Days Sint-Adm | 4.32 ± 3.39 | 3.86 ± 3.11 | 4.26 ± 3.26 | 4.46 ± 3.56 | 4.56 ± 3.74 |
| Days Adm-Dead | 5.88 ± 4.95 | 2 ± 2.34 | 6.12 ± 4.61 | 5.83 ± 5.03 | 5.82 ± 5.24 |
| Days Sint-Dead | 10.08 ± 5.47 | 7 ± 3.39 | 10.23 ± 4.95 | 9.95 ± 5.71 | 10.47 ± 5.41 |
| IMSS Services | 3634 (34.47) | 115 (19.23) | 2185 (38.74) | 1189 (31.1) | 145 (30.02) |
| ISSSTE Services | 641 (6.08) | 12 (2.01) | 274 (4.86) | 293 (7.66) | 62 (12.84) |
| SSA Services | 5108 (48.44) | 398 (66.56) | 2631 (46.65) | 1878 (49.12) | 201 (41.61) |
| Other Public Services | 415 (3.94) | 12 (2.01) | 175 (3.1) | 190 (4.97) | 38 (7.87) |
| Private Services | 746 (7.08) | 61 (10.2) | 375 (6.65) | 273 (7.14) | 37 (7.66) |

Table 2 presents the results of the comparisons between the two main outcomes. The vast majority of hospitalised individuals were men; people in this group were older, with a higher prevalence of morbidity, both alone and in combination (p≤0.01). Immuno-suppression and smoking were also more frequent than in the no hospitalised; pneumonia, intubation, and admission to the ICU were especially higher in this group (p≤0.01). Regarding mortality, a higher percentage of no survivors were men. In comparison with the survivors, no survivors were older, with a higher prevalence of morbidities alone and in combination (p≤0.01). Hospitalisation, ICU admission, and intubation were more frequent on no survivors (p≤0.01). A higher percentage of no survivors were attended in the IMSS, while survivors had been frequently attended at the SSA services (p≤0.01). For both outcomes, obesity alone and the days between the presentation of symptoms, the admission to the hospital and the death, were no different (p≥0.05).

**Table 2.** General characteristics by outcome

|  | Total<br>n = 9946 | No Hospitalised<br>n= 6024 | Hospitalised<br>n= 3922 | p value | Survivor<br>n = 8983 | No Survivor<br>n= 963 | p value |
| --- | --- | --- | --- | --- | --- | --- | --- |
| Age, years | 48.15 ± 14.35 | 44.18 ± 12.9 | 54.24 ± 14.34 | <0.0001 | 46.96 ± 13.91 | 59.26 ± 13.63 | 0.0001 |
| Sex (men) | 5753 (57.84) | 3204 (53.19) | 2549 (64.99) | 0.0001 | 5086 (56.62) | 667 (69.26) | 0.0001 |
| Hypertension | 2265 (22.98) | 938 (15.73) | 1327 (34.1) | 0.0001 | 1845 (20.74) | 420 (43.8) | 0.0001 |
| Obesity | 2053 (20.82) | 1066 (17.86) | 987 (25.35) | 0.0001 | 1764 (19.82) | 289 (30.1) | 0.0001 |
| Diabetes | 1834 (18.61) | 668 (11.2) | 1166 (29.97) | 0.0001 | 1470 (16.53) | 364 (37.92) | 0.0001 |
| Cardiovascular Disease | 310 (3.15) | 110 (1.84) | 200 (5.14) | 0.0001 | 246 (2.77) | 64 (6.68) | 0.0001 |
| Chronic kidney disease | 219 (2.22) | 54 (0.91) | 165 (4.24) | 0.0001 | 156 (1.75) | 63 (6.58) | 0.0001 |
| COPD | 270 (2.74) | 72 (1.21) | 198 (5.09) | 0.0001 | 187 (2.1) | 83 (8.65) | 0.0001 |
| Asthma | 356 (3.61) | 237 (3.98) | 119 (3.06) | 0.0177 | 325 (3.66) | 31 (3.23) | 0.4925 |
| Number of Comorbidities | 0.74 ± 0.97 | 0.52 ± 0.82 | 1.07 ± 1.08 | <0.0001 | 0.67 ± 0.92 | 1.36 ± 1.16 | <0.0001 |
| No Comorbidities | 5273 (53.64) | 3812 (64.01) | 1461 (37.7) | 0.0001 | 5018 (56.54) | 255 (26.7) | 0.0001 |
| Combinated Chronic Diseases |  |  |  |  |  |  |  |
| Diabetes & Hypertension & Obesity | 322 (3.24) | 122 (2.03) | 200 (5.1) | 0.0001 | 246 (2.74) | 76 (7.89) | 0.0001 |
| Diabetes & Hypertension | 638 (6.41) | 193 (3.2) | 445 (11.35) | 0.0001 | 485 (5.4) | 153 (15.89) | 0.0001 |
| Diabetes & Obesity | 228 (2.29) | 100 (1.66) | 128 (3.26) | 0.0001 | 190 (2.12) | 38 (3.95) | 0.0003 |
| Hypertension & Obesity | 395 (3.97) | 192 (3.19) | 203 (5.18) | 0.0001 | 333 (3.71) | 62 (6.44) | 0.0001 |
| Hypertension | 908 (9.13) | 431 (7.15) | 477 (12.16) | 0.0001 | 779 (8.67) | 129 (13.4) | 0.0001 |
| Obesity | 1103 (11.09) | 650 (10.79) | 453 (11.55) | 0.2385 | 990 (11.02) | 113 (11.73) | 0.5028 |
| Diabetes | 643 (6.46) | 253 (4.2) | 390 (9.94) | 0.0001 | 547 (6.09) | 96 (9.97) | 0.0001 |
| Pregnancy | 48 (0.48) | 34 (0.56) | 14 (0.36) | 0.1525 | 43 (0.48) | 5 (0.52) | 0.7329 |
| Immuno-suppressed | 195 (1.98) | 79 (1.32) | 116 (2.98) | 0.0001 | 149 (1.68) | 46 (4.8) | 0.0001 |
| Smoking | 894 (9.07) | 486 (8.15) | 408 (10.49) | 0.0001 | 795 (8.94) | 99 (10.33) | 0.1524 |
| Pneumonia | 2971 (29.87) | 327 (5.43) | 2644 (67.41) | 0.0001 | 2245 (24.99) | 726 (75.39) | 0.0001 |
| Hospitalized | 3922 (39.43) | -- | 3922 (100) | 0.0001 | 3054 (34.0) | 868 (90.13) | 0.0001 |
| Intubated | 460 (4.63) | -- | 460 (11.73) | 0.0001 | 234 (2.61) | 226 (23.47) | 0.0001 |
| ICU | 486 (4.89) | -- | 486 (12.39) | 0.0001 | 294 (3.27) | 192 (19.94) | 0.0001 |
| Dead | 963 (9.68) | 95 (1.58) | 868 (22.13) | 0.0001 | -- | 963 (100) | 0.0001 |
| Days Sint-Adm | 4.35 ± 3.4 | 4.31 ± 3.3 | 4.41 ± 3.55 | 0.1631 | 4.37 ± 3.38 | 4.19 ± 3.62 | 0.1107 |
| Days Adm-Dead | 5.9 ± 4.95 | 6.03 ± 5.33 | 5.89 ± 4.91 | 0.8005 | -- | 5.93 ± 4.93 | 0.8005 |
| Days Sint-Dead | 10.09 ± 5.47 | 10.79 ± 6.38 | 10.02 ± 5.36 | 0.1920 | -- | 10.12 ± 5.45 | 0.1920 |
| IMSS Services | 3519 (35.38) | 2023 (33.58) | 1496 (38.14) | 0.0001 | 3069 (34.16) | 450 (46.73) | 0.0001 |
| ISSSTE Services | 629 (6.32) | 212 (3.52) | 417 (10.63) | 0.0001 | 562 (6.26) | 67 (6.96) | 0.3945 |
| SSA Services | 4710 (47.36) | 3137 (52.08) | 1573 (40.11) | 0.0001 | 4331 (48.21) | 379 (39.36) | 0.0001 |
| Other Public Services | 403 (4.05) | 191 (3.17) | 212 (5.41) | 0.0001 | 359 (4) | 44 (4.57) | 0.3874 |
| Private Services | 685 (6.89) | 461 (7.65) | 224 (5.71) | 0.0002 | 662 (7.37) | 23 (2.39) | 0.0001 |

The full logistic regression model for hospitalisation containing seven covariates was statistically significant (n=9847; Goodness-of-fit test x^2^=1080.18, df=536, p<0.001; Area under the ROC curve=0.8886). Controlling for other predictors in the model, men were about 1.54 times as likely to be hospitalized than women (p<0.001, 95% C.I. 1.37-1.74); individuals aged 50-74 and ≥74 years were more likely to be hospitalized than people from 25-49 years (OR 2.05, p<0.001, 95% C.I. 1.81-2.32, and OR 23.84, p<0.001, 95% C.I. 2.90-5.15, respectively). People with chronic kidney disease and with COPD was 2.01 and 1.73 times more likely to be hospitalized than people without these comorbidities (p=0.001, 95% C.I. 1.33-3.04, and p=0.003, 95% C.I. 1.20-2.50, respectively). In contrast to individuals without the three main comorbidities, people with diabetes and hypertension, diabetes alone, the combination of the three, diabetes and obesity, hypertension and obesity, obesity alone, and hypertension had 2.60, 2.14, 1.85, 1.78, 1.65, 1.64, and 1.54 times the risk of being hospitalised. People attended in the ISSSTE, IMSS and other services were more likely to be hospitalised than people attended on private services (OR=3.08, 2.15, and 1.72, respectively). Pneumonia had the highest OR for being hospitalised instead of not (OR= 33.62, p<0.001, 95% C.I. 29.22-38.68) in comparison with not having pneumonia.

The full logistic regression model for mortality containing twelve covariates was statistically significant (n=9845; Goodness-of-fit test x^2^=2180.73, df=1140, p<0.001; Area under the ROC curve=0.8840). Controlling for other predictors in the model, the highest risk of death was given by IMSS in comparison with private services (OR 9.25, p<0.001, 95% C.I. 5.61-15.24); for the same variable, SSA, other and ISSSTE services had more risk of death in comparison to private services (OR=3.56, 2.45 and 2.25). Men had more risk of death in comparison to women (OR=1.53, p<0.001, 95% C.I. 1.30-1.81) and individuals aged 50-74 and ≥75 years were more likely to die than people from 25-49 years (OR 1.96, p<0.001, 95% C.I. 1.63-2.34, and OR 3.74, p<0.001, 95% C.I. 2.80-4.98, respectively). People with chronic kidney disease and with COPD had 1.44 and 1.68 times the risk to die than people without these comorbidities (p=0.047, 95% C.I. 1.01-2.06, and p=0.002, 95% C.I. 1.22-2.31, respectively). When the combination of the main chronic diseases was compared with not having these diseases, the combination of the three had the highest risk of die (OR=2.10; p<0.001, 95% C.I. 1.50-2.93), followed by diabetes and obesity, diabetes and hypertension, hypertension and obesity, obesity alone, diabetes, and hypertension, that had 2.06, 1.92, 1.88, 1.74 and 1.49 times the risk of dying. Pregnant women had 3.56 times the risk of dying in comparison with no pregnant women (p=0.025, 95% C.I. 1.17-10.80) and immuno-suppressed individuals are about 1.70 times more likely to die than people without this condition. People who developed pneumonia were 2.57 more likely to die than people without it (p<0.001, 95% C.I. 2.11-3.13), and those who had been hospitalised, intubated and admitted to ICU had a higher risk of dying than their counterparts (OR 5.02, p<0.001, 95% C.I. 3.88-6.50, OR 4.27, p<0.001, 95% C.I. 3.26-5.59, and OR 1.79, p<0.001, 95% C.I. 1.36-2.36, respectively).

## DISCUSSION

For our knowledge, this is the first study of COVID-19 reporting the risk factors for hospitalisation and death in the Mexican population. Men, individuals in the older ages groups, CKD, COPD, with chronic diseases in combination or alone, who developed pneumonia and were attended in any public health service, were at higher risk of being hospitalised than people who were not. For death, the same factors in addition to pregnancy, immuno-suppression, hospitalisation, intubation, and admission to the ICU, increased the risk when compared to the survivors.

Shortly after the COVID-19 disease had been reported as a pandemic, small case series reports of individuals attended in different hospitals in China had raised (11-13). Endpoints, especially admission to ICU and invasive ventilation, were frequently reported. Hospitalisation, one of the events of significant concern to health systems due to the risk of saturation, had not been used as a primary outcome. In fact, a large hospitalisation rate has been reported by Guan et al. (14) who informed that 93.6% of the individuals with COVID-19 received hospital care. In-hospital death mortality had been reported to be as high as 28% and 97% for individuals requiring mechanical ventilation (15). In the present analysis, 40% of the individuals were attended in hospitals. Of the 3922 individuals who had been hospitalised, 67% developed pneumonia, 88% were intubated, and 12% were admitted to the ICU. The death rate within hospitalisation was 22%.

Overall lethality of COVID-19 in the present study was 9.2%. The higher proportion of deaths was observed for the older age group (27.95%) and in individuals with any type of morbidity (73.3%).

Li et al. published a meta-analysis that aimed to analyse the clinical data, discharge rate, and fatality rate of COVID-19 patients for clinical help. Male sex was strongly related to adverse outcomes (16). In the present analysis, men also have more risk of being hospitalised and died, 54% and 53%, respectively, than women. It had been repeatedly informed that fatality rates for males are two to three times higher than for females (17). Wenham et al. (18) had suggested that gender-related social factors, immunological differences, hormonal disparities, and lifestyle habits may play a role in the sex differences for COVID-19.

Regarding age, in Europe, a higher mortality rate had been reported in older age groups (19). A similar pattern has been reported for China (20). It could be thought that in Mexico the risk for the older population would be lower, as the ageing group is smaller compared to these countries. Notwithstanding the hospitalisation and mortality risks were more than twice in the older age groups in our analysis. The reduced immune response and the increased prevalence of multimorbidity that characterised this age group can explain the higher risk of both outcomes.

In the same line, morbidity increased the risk of hospitalisation and death in the present analysis, mainly when chronic degenerative diseases occur simultaneously. From individuals requiring hospitalisation 62% had comorbidities, primarily hypertension (34%), diabetes (30%), and obesity (25%). In fact, the presence of these three diseases in the same person increases 85% of the risk of hospitalisation. For mortality, a similar pattern can be observed. In individuals who died, hypertension was present in 44%, diabetes in 38%, and obesity in 30%. The risk of dying in individuals presenting the combination of these three diseases was 2.10 times the risk compared with those without these diseases. It is important to note that individuals who died with no morbidities (27%) were younger (56.17±13.42 years) than those who died and did have morbidities (60.41±13.59 years). Considering that in Mexico it had been reported a high proportion of undiagnosed chronic diseases (21), it can be hypothesized that a proportion of hospitalisations and deaths from COVID-19 can be related to undiagnosed morbidities.

Data on pregnancy and COVID-19 is limited, but based on the experience with influenza, SARS, and MERS, pregnant women, especially those in the second and third trimesters of gestation, have a higher risk of complications and death in comparison with no pregnant women (22-24). In the present analysis pregnant women, in comparison to no pregnant, were 3.56 times more likely to die because of coronavirus. It has to be noted that 80% of pregnant women who died, also had hypertension, obesity, or diabetes (data not shown).

Hospitalisation in this population seems to act as a strong risk factor for dying, and this risk further increased when individuals were admitted to the ICU and were intubated. This association can be related to the fact that the vast majority of people with COVID-19 who accessed health services are complicated cases of COVID-19.

Concerning health services, it was observed that the risk of dying is more than twice in public services than in private ones. Of especial interest is the case of IMSS that had 8.25 times the risk of dying. Public hospitals in Mexico are the health services with the greatest demand, as are more affordable and accessible for the vast majority of the Mexican population. This situation puts these institutions in the risk of over-limit their response capacity, increasing the severity and death rates associated with the health services saturation. This raises the issue of inequality that hinders access to quality care, late arrivals, overcrowded services and poor staffing in public hospital vs. world class care in the private sector.

This study has some limitations. First, given the dynamics of the disease and that in Mexico there are insufficient resources to apply the tests massively, the rates were calculated based on sentinel information that includes all deaths, but not all mild (asymptomatic) or moderate cases that did not access to health services, so the prevalence of the disease could be underestimated and lethality could be overestimated. Therefore, when interpreting the data, it would be convenient to take into account the estimation of real cases, which are mostly mild cases of the disease. Second, more detailed patient information, particularly, dates of hospital discharge of patients who do not die, was unavailable at the time of analysis; this information would be of vital importance to assess the possible saturation of health services and to assess the use of resources.

Nonetheless, this study is, to our knowledge, the largest case series to date of COVID-19 in Mexico, with 10,544 individuals from all over the country, and provides further information on the clinical and epidemiological features of patients. It presents the latest status of COVID-19 in Mexico and a wide range of clinical manifestations can be seen and are associated with adverse outcomes.

## CONCLUSIONS

COVID-19 placed substantial strains on the health systems worldwide. For countries starting the accelerated contagion phase, it is important to identify individuals with poor prognosis at an early stage in order to better allocated limited resources. In this respect, the present study points out that in Mexico, where an important proportion of the population develops two or more chronic conditions simultaneously, high mortality is a sever outcome for those infected by SARS-CoV-2.

## Data Availability

Data can be found from the Mexican Ministry of Health (Secretaria de Salud, SS)
We downloaded the data available on 23rd. April 2020

https://datos.gob.mx/busca/dataset/informacion-referente-a-casos-covid-19-en-mexico

## ACKNOWLEDGMENTS

Does not apply.

---

**Table 3.** Logistic regression model for the risk of hospitalisation

|  | Odds Ratio | Std. Err. | z | p>z | 95% C.I. |  |
| --- | --- | --- | --- | --- | --- | --- |
| Men | 1.54 | 0.09 | 7.32 | <0.001 | 1.37 | 1.74 |
| Age 25-49 years (reference) |  |  |  |  |  |  |
| Age 50-74 years | 2.05 | 0.13 | 11.31 | <0.001 | 1.81 | 2.32 |
| Age ≥ 75 years | 3.84 | 0.56 | 9.32 | <0.001 | 2.90 | 5.10 |
| CKD | 2.01 | 0.42 | 3.33 | 0.001 | 1.33 | 3.04 |
| COPD | 1.73 | 0.32 | 2.96 | 0.003 | 1.20 | 2.50 |
| None (reference) |  |  |  |  |  |  |
| Diabetes & Hypertension & Obesity | 1.85 | 0.31 | 3.74 | <0.001 | 1.34 | 2.56 |
| Diabetes & Hypertension | 2.60 | 0.32 | 7.73 | <0.001 | 2.04 | 3.31 |
| Diabetes & Obesity | 1.78 | 0.34 | 3.01 | 0.003 | 1.22 | 2.59 |
| Hypertension & Obesity | 1.65 | 0.24 | 3.45 | 0.001 | 1.24 | 2.19 |
| Hypertension | 1.54 | 0.16 | 4.2 | <0.001 | 1.26 | 1.88 |
| Obesity | 1.64 | 0.15 | 5.41 | <0.001 | 1.37 | 1.95 |
| Diabetes | 2.14 | 0.25 | 6.43 | <0.001 | 1.70 | 2.69 |
| Pneumonia | 33.62 | 2.40 | 49.14 | <0.001 | 29.22 | 38.68 |
| Private Services (reference) |  |  |  |  |  |  |
| IMSS Services | 2.15 | 0.28 | 5.85 | <0.001 | 1.67 | 2.78 |
| ISSSTE Services | 3.08 | 0.52 | 6.68 | <0.001 | 2.21 | 4.28 |
| SSA Services | 0.82 | 0.11 | -1.56 | 0.118 | 0.63 | 1.05 |
| Other Services | 1.72 | 0.33 | 2.88 | 0.004 | 1.19 | 2.49 |
| Constant | 0.03 | 0.01 | -14.61 | <0.001 | 0.02 | 0.04 |

**Table 4.** Logistic regression model for the risk of death

|  | Odds Ratio | Std. Err. | z | p>z | 95% C.I. |  |
| --- | --- | --- | --- | --- | --- | --- |
| Men | 1.53 | 0.13 | 5.01 | <0.001 | 1.30 | 1.81 |
| Age 25-49 years (reference) |  |  |  |  |  |  |
| Age 50-74 years | 1.96 | 0.18 | 7.29 | <0.001 | 1.63 | 2.34 |
| Age ≥ 75 years | 3.74 | 0.55 | 9.01 | <0.001 | 2.80 | 4.98 |
| CKD | 1.44 | 0.26 | 1.99 | 0.047 | 1.01 | 2.06 |
| COPD | 1.68 | 0.27 | 3.16 | 0.002 | 1.22 | 2.31 |
| None (reference) |  |  |  |  |  |  |
| Diabetes & Hypertension & Obesity | 2.10 | 0.36 | 4.32 | <0.001 | 1.50 | 2.93 |
| Diabetes & Hypertension | 1.92 | 0.25 | 4.94 | <0.001 | 1.48 | 2.49 |
| Diabetes & Obesity | 2.06 | 0.44 | 3.39 | 0.001 | 1.35 | 3.12 |
| Hypertension & Obesity | 1.88 | 0.33 | 3.58 | <0.001 | 1.33 | 2.65 |
| Hypertension | 1.49 | 0.19 | 3.04 | 0.002 | 1.15 | 1.92 |
| Obesity | 1.74 | 0.23 | 4.21 | <0.001 | 1.35 | 2.26 |
| Diabetes | 1.50 | 0.21 | 2.81 | 0.005 | 1.13 | 1.98 |
| Pregnancy | 3.56 | 2.01 | 2.24 | 0.025 | 1.17 | 10.80 |
| Inmuno-suppressed | 1.70 | 0.35 | 2.57 | 0.010 | 1.13 | 2.55 |
| Pneumonia | 2.57 | 0.26 | 9.45 | <0.001 | 2.11 | 3.13 |
| Hospitalized | 5.02 | 0.66 | 12.25 | <0.001 | 3.88 | 6.50 |
| Intubated | 4.27 | 0.59 | 10.51 | <0.001 | 3.26 | 5.59 |
| ICU | 1.79 | 0.25 | 4.18 | <0.001 | 1.36 | 2.36 |
| Private Services (reference) |  |  |  |  |  |  |
| IMSS Services | 9.25 | 2.36 | 8.72 | <0.001 | 5.61 | 15.24 |
| ISSSTE Services | 2.25 | 0.64 | 2.86 | 0.004 | 1.29 | 3.91 |
| SSA Services | 3.56 | 0.89 | 5.08 | <0.001 | 2.18 | 5.81 |
| Other Services | 2.45 | 0.75 | 2.93 | 0.003 | 1.34 | 4.46 |
| Constant | 0.00 | 0.00 | 17.18 | <0.001 | 0.00 | 0.00 |

## Notes

### Competing Interest Statement

The authors have declared no competing interest.

### Funding Statement

This project was supported by a grant from the Secretaria de Educacion, Ciencia, Tecnologia e Innovacion de la Ciudad de Mexico CM-SECTEI/041/2020 “Red colaborativa de Investigación Traslacional para el Envejecimiento Saludable de la Ciudad de Mexico” (RECITES).
The funder had no role in study design, data collection and analysis, decision to publish, or preparation of the manuscript.

## REFERENCES

1. World Health Organization. Situation report - 90. Coronavirus disease 2019 (COVID-19). 2020.

2. Panamerican Health Organization. COVID-19 - Respuesta de la OPS/OMS Reporte 2. 2020.

3. Secretaría de Salud. Noticias, comunicados, conferencias México2020 [Available from: https://coronavirus.gob.mx/noticias/.

4. Li Q. GX, Wu P., Wang X., Zhou L., Tong Y.. Early transmission dynamics in Wuhan, China, of novel coronavirus-infected pneumonia. N Engl J Med. 2020;382 1199-207.

5. Chen N, Zhou M, Dong X, Qu J, Gong F, Han Y, et al. Epidemiological and clinical characteristics of 99 cases of 2019 novel coronavirus pneumonia in Wuhan, China: a descriptive study. Lancet. 2020.

6. Ren LL, Wang YM, Wu ZQ, Xiang ZC, Guo L, Xu T. Identification of a novel coronavirus causing severe pneumonia in human: a descriptive study. Chin Med J. 2020.

7. Huang C, Wang Y, Li X, Ren L, Zhao J, Hu Y. Clinical features of patients infected with 2019 novel coronavirus in Wuhan, China. Lancet. 2020;395 (10223):497–506.

8. Wang T, Du Z, Zhu F, Cao Z, An Y, Gao Y, et al. Comorbidities and multi-organ injuries in the treatment of COVID-19. Lancet. 2020;395(e52).

9. Parra-Rodríguez P, González-Meljem JM, Gómez-Dantés H, Gutiérrez-Robledo H, López-Ortega M, García-Peña C, et al. The Burden of Disease in Mexican Older Adults: Premature Mortality Challenging a Limited-Resource Health System. J Aging Health. 2019;27.

10. Secretaría de Salud. Bases de datos COVID-19. 2020.

11. Cai J, Xu J, Lin D, Yang Z, Xu L, Qu Z, et al. A Case Series of children with 2019 novel coronavirus infection: clinical and epidemiological features. Clin Infect Dis. 2020.

12. Wang D, Yin Y, Hu C, Liu X, Zhang X, Zhou S, et al. Clinical course and outcome of 107 patients infected with the novel coronavirus, SARS-CoV-2, discharged from two hospitals in Wuhan, China. Crit Care. 2020;24(1):188.

13. Chan JF-W, Yuan S, Kok K-H, To KK-W, Chu H, Yang J, et al. A familial cluster of pneumonia associated with the 2019 novel coronavirus indicating person-to-person transmission: a study of a family cluster. Lancet. 2020.

14. Guan W-J, Ni Z-Y, Hu Y, Liang W-H, Ou C-Q, He J-X, et al. Clinical characteristics of coronavirus disease 2019 in China. N Engl J Med. 2020.

15. Zhou F, Yu T, Du R, Fan G, Liu Y, Liu Z, et al. Clinical course and risk factors for mortality of adult inpatients with COVID-19 in Wuhan, China: a retrospective cohort study. Lancet. 2020.

16. Li LQ, Huang T, Wang YQ, Wang ZP, Liang Y, Huang TB, et al. cOvID-19 patients' clinical characteristics, discharge rate, and fatality rate of meta-analysis. J Med Virol. 2020.

17. Porcheddu R, Serra C, Kelvin D, Kelvin N, Rubino S. Similarity in case fatality rates (CFR) of COVID-19/SARS-COV-2 in Italy and China. J Infect Dev Ctries. 2020;14:125–8.

18. Wenham C, Smith J, Morgan R, Gender Group C-W. COVID-19: the gendered impacts of the outbreak. Lancet. 2020;395:846–8.

19. Lippi G, Mattiuzzi C, Sanchis-Gomar F, Henry BM. Clinical and Demographic Characteristics of Patients Dying From COVID-19 in Italy Versus China. J Med Virol. 2020.

20. Wang W, Tang J, Wei F. Updated understanding of the outbreak of 2019 novel coronavirus (2019-nCoV) in Wuhan, China. J Med Virol. 2020;92:441–7.

21. Rojas-Martínez R, Aguilar-Salinas CA, Jiménez-Corona A, Gómez-Pérez FJ, Barquera S, Lazcano-Ponce E. Prevalence of obesity and metabolic syndrome components in Mexican adults without type 2 diabetes or hypertension. Salud Pública Mex. 2020;54(1):7–12.

22. Donders F, Lonnée-Hoffmann R, Tsiakalos A, Mendling W, Martinez de Oliveira J, Judlin P, et al. ISIDOG COVID-19 Guideline Workgroup. ISIDOG Recommendations Concerning COVID-19 and Pregnancy. Diagnostics. 2020;10:243.

23. Lam CM, Wong SF, Leung TN, Chow KM, Yu WC, Wong TY, et al. A case-controlled study comparing clinical course and outcomes of pregnant and non-pregnant women with severe acute respiratory syndrome. BJOG. 2004;111: 771–4.

24. Jamieson DJ, Theiler RN, Rasmussen SA. Emerging infections and pregnancy. Emerg Infect Dis. 2006;12:1638–43.

